# How Developmental Disorders Changed Before and After COVID-19 Pandemic

**DOI:** 10.1101/2025.08.01.25332582

**Authors:** Derek V Pierce, Yipeng Song, Yang S. Liu, Fernanda Talarico, Yutong Li, Julie Tian, Jianshan Chen, Mengzhe Wang, Bo Cao

## Abstract

The COVID-19 pandemic has had a lasting impact on mental health, with lingering effects on the healthcare utilization of developmental disorders, such as autism spectrum disorder, attention-deficit/hyperactivity disorder, and intellectual disabilities. This study explores changes in developmental disorder healthcare utilization in the Alberta healthcare system before and after the pandemic. Results indicate an overall increase in the healthcare utilization of developmental disorders from 2018 to 2022. The ongoing impact of the pandemic on developmental disorders highlights the value of better surveillance, mental health support, and informed policy decisions to ensure individuals with developmental disorders and their families receive the necessary support and resources.

## Introduction

The coronavirus disease 2019 (COVID-19) pandemic has had a significant impact on people’s physical and mental health (1,2), affecting people of all ages (3,4), but young individuals seem to be the most affected (5,6). While much of the initial impact has subsided, lingering mental health issues remain (7,8), especially for individuals with developmental disorders (9). A concerning development is the increased utilization of the healthcare system for developmental disorders (10), such as autism spectrum disorder (ASD), attention-deficit/hyperactivity disorder (ADHD) (11), and intellectual disabilities.

Developmental disorders are generally more chronic in nature and less likely to resolve over time (12). The impact on a younger population in need of specialized support, such as school aides and therapists, will have ongoing impacts as these individuals age (13). This trend raises concerns about the potential for increased costs to the healthcare system in the future and adds urgency to addressing this issue.

The sudden shift to remote learning and increased social isolation during COVID-19 may have been an aggravating factor for individuals with developmental disorders and their families (14). These changes could have potentially exacerbated their symptoms and made it more difficult for their long-term success. The disruption of critical resources may have impacted the ability of students with developmental disorders to access necessary support. While previous studies have shown the significant impact of the pandemic on mental health, leading to increased rates of various disorders such as depression, anxiety, and stress (14), there remains limited insight on developmental disorders. This highlights the need to better understand how developmental disorders’ prevalence and presentation changes before and after the pandemic as well as factors driving those changes. The Canadian healthcare system offers universal coverage for the population. Alberta, a province of over 4 million residents provides a representative sample for investigating population level healthcare utilization. This study seeks to examine the changes in developmental disorder healthcare utilization in Alberta, Canada, before and after the COVID-19 pandemic.

## Methods

This research is part of a collaborative effort under an interchange agreement, focused on addressing health data priorities and providing recommendations to the Ministries in Alberta, Canada. This study was approved by the Research Ethics Boards, University of Alberta (Pro00072946). Informed consent was waived due to the minimal risks involved in the secondary analysis of anonymized data. The authors had no access to identify individual participants during and after data collection.

### Data Source

This study used the administrative healthcare databases of practitioner claims (CLM), emergency department visits (ED), and inpatient services (INP) in the Ministry of Health in Alberta. The data was accessed from April 1^st^ 2021 to Feb 2^nd^ 2023. This period was selected to capture both the immediate and evolving effects of the pandemic on healthcare utilization.

### Cohort

The study cohort included all residents of Alberta with active health insurance coverage for public healthcare services. Exclusion criteria included individuals with non-active health insurance during the study period. This cohort was dynamic in nature, as individuals may enter or exit the cohort annually.

### Outcome

Mental health-related diagnoses were selected based on the International Classification of Diseases, 9th or 10th revision (ICD-9 and ICD-10), available in the records from physician’s office visits, emergency department visits, and inpatient services. Developmental disorder related services were identified from records with a primary diagnosis code of the following mental disorders: ADHD, ASD, intellectual disability, communication disorder, special learning disorder, coordination disorder, tics, and conduct disorder, The case definitions for each disorder are provided in Table S1.

### Data processing

We extracted the number of unique patients and utilization events related to mental health concerns between 2016 and 2022. To normalize the data, for each year, we calculated the service utilization by calculating the proportion of unique patients with developmental disorders health care utilization, per 1000 patients with active Alberta Health care insurance. The result is the number of unique patients related to developmental disorders per 1000 patients for each year.

To understand the impact of the COVID-19 pandemic on health services utilization, we estimated the expected utilization numbers and the associated standard deviations for 2020, 2021, and 2022, assuming a linear trend in utilization from 2016 to 2019. We also explored the observed annual temporal trend of utilization as a comparison.

For the monthly temporal trend, we extracted the number of unique patients with developmental disorder related services for each month from January 2016 to December 2022. Similar to the yearly data, these utilization numbers were adjusted by the population size of patients with active Alberta health care insurance with health care utilization, in the corresponding month of the year.

## Results

The data collected in this study consists of the healthcare utilization of developmental disorders per 1000 patients in four groups (All, CLM, ED, and INP) from 2016 to 2022. The data was analyzed to identify various trends in the data, including quarterly, seasonal, and annual trends.

**Table 1.** The cohort size reported in this table is derived from the sum of unique records in the system spanning from 2016 to 2022. “Developmental” represents the subjects with any health care utilization record of developmental disorders during this period. “General population” refers to the population encompassing all other individuals without developmental disorders.

| Cohort | Sample Size | Mean age | Female (%) |
| --- | --- | --- | --- |
| Developmental | 387,337 | 26.2 | 45 |
| General population | 4,960,783 | 39.6 | 50 |

Figure 1 illustrates the observed results compared to an expected linear growth pattern based on the changes observed between 2016 and 2019. While the expected growth pattern follows a consistent upward trajectory, the actual observed results deviate from this trend, particularly when considering all sources combined. This is contrasted with the sharp dip in ED visits in 2020 when compared to previous ED trends and observations from the other data.

**Figure 1.**
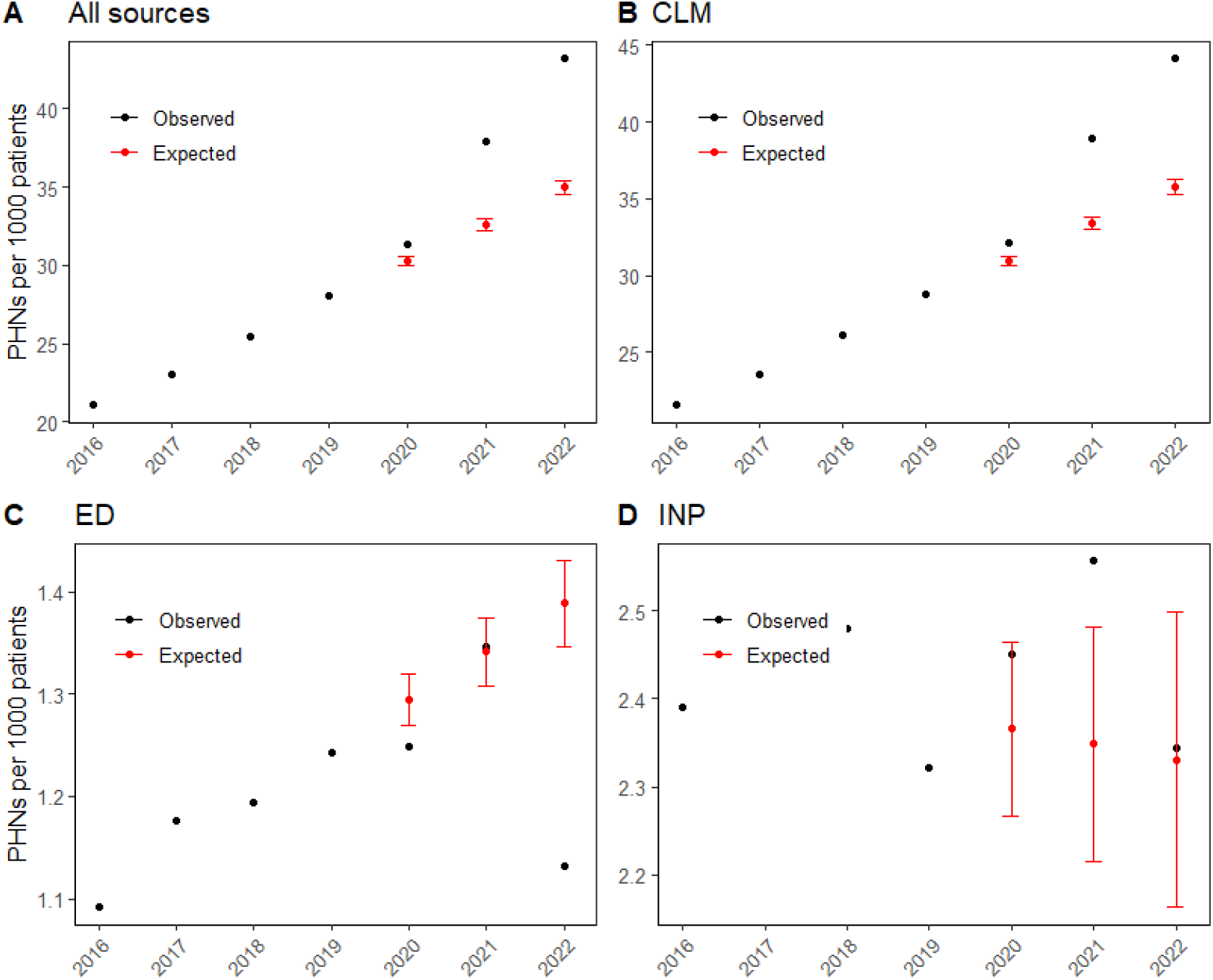
Trend of the observed and expected number of patients with development disorders scaled for 1000 patients along different years. ‘All sources’ is the combination of physician’s office visits (CLM), emergency department (ED) visits, and inpatient (INP) data. PHN is the Personal Health Number associated with unique individuals.

An increasing trend in the healthcare utilization of developmental disorders was observed for the combination of all sources and CLM, while a decrease in ED visits and INP was seen over the five-year period (Figure 2). The overall healthcare utilization in the ‘All’ group increased from 23.3 patients per 1000 patients in the first quarter (Q1) of 2018 to 41.6 in 2022 quarter 3 (Q3), indicating an overall increase of 78.5%. The CLM group experienced a similar increase, from 24.1 patients per 1000 patients in 2018-Q1 to 43.5 in 2022-Q3, marking an 80.5% increase. The ED group’s healthcare utilization rose from 1.06 patients per 1000 patients in 2018-Q1 to 0.84 in 2022-Q3, reflecting a decrease of 20.8%. The INP group saw an increase from 2.73 patients per 1000 patients in 2018-Q1 to 2.03 in 2022-Q3, representing a 25.6% decrease (Figure 2).

**Figure 2:**
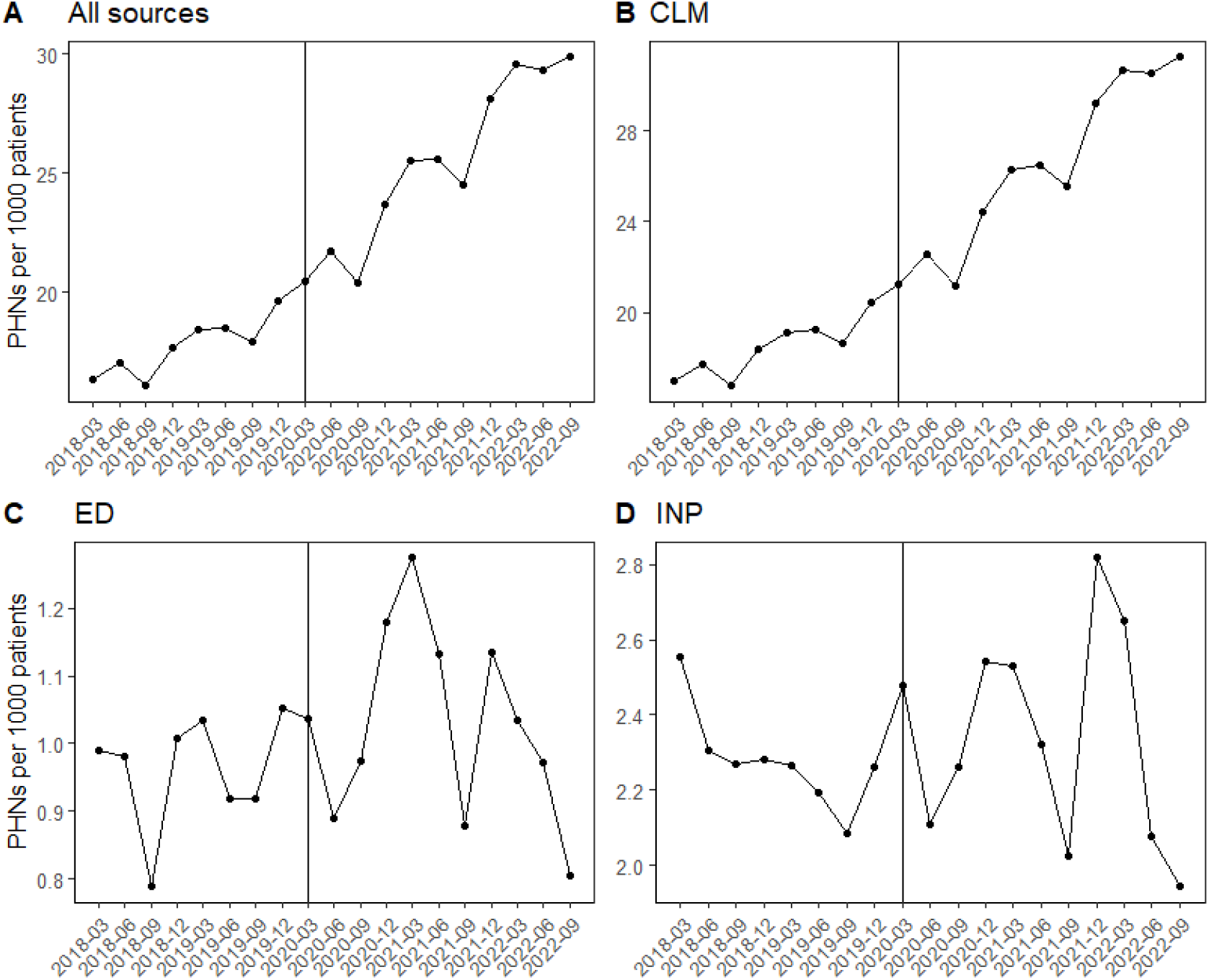
The number of patients with development disorders scaled for 1000 patients along different quarters. ‘All sources’ is the combination of physician’s office visits (CLM), emergency department (ED) visits, and inpatient (INP) data.

There were fluctuations in the healthcare utilization of developmental disorders within the groups when comparing quarters. In general, the ‘All’ and CLM groups experienced an increase in healthcare utilization in the second and fourth quarters of each year (Figure 2). The ED group showed variability across quarters, with a notable decrease in healthcare utilization in 2022-Q3 (0.84) compared to 2018-Q1 (1.06). The INP group exhibited a general decrease in healthcare utilization over time, with the lowest rate in 2022-Q3 (2.03).

When it comes to trends on a seasonal basis, the ‘All’ and CLM groups displayed a higher healthcare utilization of developmental disorders in the summer (second quarter) and winter (fourth quarter) seasons (Figure 2). This pattern does not seem to be influenced by the COVID-19 pandemic (Figure 2). In contrast, the ED group showed no clear seasonal pattern, while the INP group demonstrated a decline in healthcare utilization throughout the study period, with the lowest healthcare utilization in the fall (third quarter) of 2022 (Figure 2).

Other trends of interest include a steep annual increase in the healthcare utilization of developmental disorders for the ‘All’ group between 2020-Q4 (36.9) and 2021-Q1 (40.5), with an increase of 3.6. Similarly, the CLM group experienced its steepest annual increase between 2020-Q4 (37.9) and 2021-Q1 (41.6), with an increase of 3.7; The largest quarterly change in the ‘All’ group occurred between 2020-Q2 (33.9) and 2020-Q3 (30.8), with a decrease of 3.1. The CLM group experienced its largest quarterly change between 2020-Q2 (35.1) and 2020-Q3 (31.9), with a decrease of 3.2. The ED group’s largest quarterly change took place between 2020-Q4 (1.29) and 2021-Q1 (1.37), with an increase of 0.08. For the INP group, the largest quarterly change was between 2021-Q4 (3.06) and 2022-Q1 (2.74), with a decrease of 0.32. Conversely, there were also periods of stability in the healthcare utilization of developmental disorders in the ED group between 2018-Q1 (1.06) and 2018-Q2 (1.06), and 2020-Q2 (1) and 2020-Q3 (1.05), with only slight changes during these periods.

Our results indicate an overall increase in the healthcare utilization of developmental disorders in the ‘All’ and CLM groups from 2018 to 2022. The ED group displayed a decrease in healthcare utilization, while the INP group demonstrated a decline over time. Seasonal trends were evident in the ‘All’ and CLM groups, with higher healthcare utilization in the summer and winter seasons. These results highlight an opportunity to enhance existing policies and support systems.

## Discussion

The result of our study provides evidence that the events surrounding the COVID-19 pandemic have had a profound and sustained impact on utilization rates in the Alberta healthcare system for developmental disorders. Developmental disorders, such as ASD and ADHD, predominantly affect children and adolescents, impacting their cognitive, emotional, and social development.

The escalation in healthcare utilization for these disorders during the pandemic suggests an increase in symptom severity, possibly due to disrupted care routines and reduced access to therapeutic and educational supports. This period of isolation and change could exacerbate existing conditions, emphasizing the need for healthcare systems to enhance their capacity to deliver services remotely and flexibly during crises. While previous research has shown the pandemic has impacted mental health (15), developmental disorder rates remain high relative to other disorders that have reduced to a utilization rate closer to their pre-COVID norms. Given the chronic nature of these conditions and the young age of the population affected, we believe it is crucial to explore this further. This underlines the importance of targeted policy interventions and the adaptation of service delivery models to ensure continuous care for vulnerable populations, especially during global health emergencies.

Several factors may have contributed to this increase in developmental disorders. The sudden shift to remote learning and increased social isolation significantly disrupted the daily routines of everyone (16,17); this included many of the support systems individuals and their families relied on for those with special needs (18). Additionally, the reduced mobility of auxiliary support staff in schools made it more difficult for these individuals to access the support they needed to succeed academically and personally during the COVID-19 lockdowns.

Our research findings emphasize several policy recommendations for the province of Alberta, which aim to address the unique challenges faced by individuals with developmental disorders during and after the COVID-19 pandemic. We suggest that enhancing the accessibility and availability of virtual support services is of value in providing continuity of care during times of social isolation or remote learning, ensuring consistent support and treatment for this vulnerable population (19). Moreover, implementing a comprehensive reintegration plan could facilitate a transition back into in-person learning environments and community-based support services (20). Such a plan could contribute to the continued academic and personal growth of individuals with developmental disorders by addressing their specific needs adequately. This also could help mitigate the longer-term costs associated with support for those who don’t successfully reintegrate into the community. It is important to acknowledge that when the burden of support primarily falls on the families themselves, it often detracts from the province’s available pool of labour and talent.

Finally, we recommend the allocation of additional funding to support research and the development of targeted interventions (21). This investment could foster a deeper understanding of the factors contributing to the observed trends and enable the mitigation of the long-term impacts of the pandemic on this at-risk population. Further research is important to comprehend the factors contributing to these trends and to develop targeted interventions addressing developmental disorders within these specific groups. By adopting any of these policy recommendations, we hope to better support individuals with developmental disorders and their families in the province of Alberta, and those beyond Alberta, Canada.

## Limitations

While our study provides valuable insights into the impact of the COVID-19 pandemic on developmental disorder utilization rates in Alberta, it is important to acknowledge some limitations. Firstly, our study is limited to Alberta’s healthcare system, and generalizing its findings to other regions or countries requires caution. Secondly, while we used administrative healthcare data to identify mental health-related diagnoses, our study is reliant on the accuracy and completeness of the coding of these diagnoses in the administrative databases. Thirdly, our study does not account for potential confounding factors, such as changes in diagnostic criteria or changes in access to healthcare services. Fourthly, as of the date of this study (January 2023), data for certain months in 2022 may not be entirely complete or representative. This is because the most recent months’ data are more susceptible to modifications, as they are still subject to updates, corrections, and additional information being added. Lastly, while our study highlights the increased utilization rates of developmental disorders, it is not able to provide a detailed understanding of the factors contributing to the observed trends. Further research is necessary to understand the underlying causes of the increased utilization rates and to identify appropriate interventions to address these trends.

## Conclusion

Overall, this study provides important insights into the impact of the COVID-19 pandemic on individuals with developmental disorders and highlights the value of continued research and action to address this issue. By improving our understanding of the pandemic’s impact on developmental disorders, we can work to ensure that individuals with these conditions receive the support and treatment they need to lead fulfilling and productive lives. We hope that the findings will increase awareness of these more chronic health issues and the need for better surveillance and mental health support for individuals with developmental disorders, especially in the context of the ongoing challenges this group will face and the added burden of costs, beyond Canada.

## Acknowledgments

This research was undertaken, in part, thanks to funding from the Canada Research Chairs program (BC), Alberta Innovates (BC and FT), Mental Health Foundation (BC), MITACS Accelerate program (BC and YSL), BBRF Young Investigator Grant from Brain & Behavior Foundation (YSL), Simon & Martina Sochatsky Fund for Mental Health (BC), Howard Berger Memorial Schizophrenia Research Fund (BC), the Abraham & Freda Berger Memorial Endowment Fund (BC), the Alberta Synergies in Alzheimer’s and Related Disorders (SynAD) program (BC, FT, YSL), University Hospital Foundation (BC) and University of Alberta (BC). The funders had no role in study design, data collection and analysis, decision to publish, or preparation of the manuscript.

## Disclaimer

This study is based in part on data provided by Alberta Health. The interpretation and conclusions contained herein are those of the researchers and do not necessarily represent the views of the Government of Alberta. Neither the Government nor Alberta Health expresses any opinion about this study.

## Data Availability Statement

Due to privacy policy restrictions individualized data cannot be shared. Data can be accessed with permission from the Ministry of Health in Alberta, Canada (https://www.alberta.ca/health-research).

## Authors’ Contributions

DVP, YS, YSL, and FT participated in the formal analysis and writing the original draft. YL, JT, JC, MW, and BC participated in conceptualization, results interpretation, draft review & editing.

